# Predictors of One-Year Mortality among Patients with Heart Failure with Preserved Ejection Fraction

**DOI:** 10.64898/2025.12.17.25342526

**Authors:** Fares Alahdab, Jack Lopuszynski, Mohammad Alkhateeb, Christopher Scott, Maliha Zahid

**Affiliations:** Department of Biomedical Informatics, Biostatistics, & Medical Epidemiology, and Department of Medicine, Division of Cardiovascular Medicine, University of Missouri, Columbia, MO; Center for Women’s Health and Reproductive Medicine, Perelman School of Medicine, University of Pennsylvania, Philadelphia, PA; Department of Medicine, University of Missouri, Columbia, MO; Department of Biostatistics, Mayo Clinic, Rochester, MN; Department of Cardiovascular Medicine, Mayo Clinic, Rochester, MN

**Keywords:** HFPEF, Albumin, nt-proBNP, Prognosis, Mortality, Risk Assessment

## Abstract

**Background:** Heart failure with preserved ejection fraction (HFpEF) is increasingly common. While widely used prognostic scores often rely on limited variables and linear assumptions, they are likely to miss complex risk patterns. We aimed to develop and internally validate prediction models for 1-year all-cause mortality after first hospitalization for decompensated HFpEF and to compare them with standard survival models.

**Methods:** We performed a retrospective cohort study (2010–2020) using electronic medical records from a large academic health system. Adults with HFpEF (EF≥50%) admitted for a first time heart failure exacerbation were included. Variables spanned demographics, comorbidities, laboratory tests, echocardiography, discharge medications, and outcomes. Data were split into training (80%) and test (20%) sets with stratification by outcome. Missing values were handled with multiple imputation by chained equations. Two tree-based classifiers (Extreme Gradient Boosting and Light Gradient Boosting) were tuned with cross-validation and evaluated by area under the receiver operating characteristic curve (AUROC) and calibration. Time-to-event models included Cox proportional hazards, random survival forest (RSF), and gradient boosting survival (GBS) with concordance index and calibration assessment. Global and local (patient-level) explainability was extracted from each model, with cross-model predictor ranking and comparison.

**Results:** We analyzed 7,840 index admissions; mean age was 78 years with 55.6% women. One-year mortality was 31.5%. Test-set AUROC was 0.751 for Extreme Gradient Boosting and 0.749 for Light Gradient Boosting with acceptable calibration. GBS achieved the highest concordance index (0.718), followed by RSF (0.711) and Cox (0.704). Lower serum albumin, older age, higher N-terminal pro B-type natriuretic peptide, renal dysfunction, and lower hemoglobin were the most consistent risk signals.

**Conclusions:** Diverse modeling families produced similar discrimination and coherent predictors. A transparent risk tool using routinely available admission data appears feasible, allowing for patient-level risk assessment for precision health.

## INTRODUCTION

Despite the ever-improving medical landscape, the prevalence of congestive heart failure (HF) continues to rise in the US. As of 2023, 6.7 million Americans have been diagnosed with HF, a number which is expected to rise to 8.5 million by 2030 (1). Currently, 33% of Americans are at risk of HF, and 1 in 4 Americans will develop the syndrome in their lifetime. While multiple phenotypes of HF exist, including heart failure with reduced ejection fraction (HFrEF), and heart failure with mildly reduced ejection fraction (HFmrEF), it is heart failure with preserved ejection fraction (HFpEF) which has been steadily increasing in prevalence, largely due to an increase in the prevalence of risk factors such as obesity, hypertension, and diabetes, and will likely eventually become the dominant phenotype (1). It is characterized by metabolic dysfunction and inflammation, leading to fibrosis and an inability of the left ventricle to relax.

Despite its increasing incidence, HFpEF, has few therapies targeting the underlying pathophysiology of this highly prevalent disease (2). Treatment for HFpEF centers around managing symptoms, including diuretics to treat congestion, or selectively treating comorbidities such as atrial fibrillation (AF) and coronary artery disease (CAD), and lifestyle changes such as weight loss that have shown efficacy in HFpEF treatment (3). Therefore, as treatments for HFpEF continue to be developed, understanding the major risk factors and prognosticators for this syndrome is of the utmost importance both in designing future clinical trials of novel therapies and in the decision-making process during clinical treatment. Traditional risk models and clinical scores such as MAGGIC and the Seattle Heart Failure Model incorporate a limited set of clinical variables and rely on linear assumptions that may not accurately reflect the prognosis in HFpEF, as recent advances in biomarker research underscore the prognostic significance of a broader range of molecules, including markers of myocardial fibrosis and renal dysfunction (4–8). In parallel, capturing nonlinear relationships and handling multidimensional data is a frequent challenge with most risk assessment and prognostication models, particularly for a complex syndrome such as HFpEF (4,9). In this study, we use machine learning techniques for risk assessment in HFpEF patients, focusing on 1-year mortality, accounting for higher-dimensional data and nonlinear relationships in a large cohort of HFpEF patients.

## METHODS

This retrospective cohort study was conducted and reported in accordance with the Strengthening the Reporting of Observational Studies in Epidemiology (STROBE) guideline (10). The machine learning models’ development and evaluation were conducted and reported in adherence to the Transparent Reporting of a multivariable prediction model for Individual Prognosis or Diagnosis (TRIPOD+AI) guideline (11).

### Study Design and Cohort Selection

We conducted a retrospective cohort study to develop and validate a machine learning model for predicting one-year all-cause mortality among patients hospitalized for decompensated HFpEF. We queried the Mayo Clinic electronic medical records (EMR) from all three sites (Rochester, MN, Jacksonville, FL, Phonex/Scottsdale, AZ) for all adult patients (age ≥18 years) admitted between January 2010 and December 2020 with a primary diagnosis of heart failure exacerbation in the setting of HFpEF, defined as a left ventricular ejection fraction (LVEF) of >50% on echocardiography performed within six months prior to admission or up to seven days following discharge. The study was approved by the Mayo Clinic Institutional Review Board, with a waiver of informed consent due to its retrospective nature and use of de-identified data.

### Data Collection and Variables

Comprehensive clinical data were extracted from the EMR for each patient. Variables included baseline demographics at time of index hospitalization, comorbidities, laboratory values at admission (including serum albumin measured on the day of admission or within seven days, NT-proBNP, high-sensitivity cardiac Troponin-T, renal function markers, and standard chemistry and hematology panels), echocardiographic parameters (including biatrial size, biventricular size and function, as well as estimated right ventricular systolic pressures (RVSP) using the modified Bernoulli equation, and estimates of right atrial pressure from inferior vena caval dimensions and respiratory collapsibility), medications at discharge, and clinical outcomes including repeat hospitalization, time to repeat hospitalizations, and one-year all-cause mortality.

### Statistical Analysis

Patients’ data were summarized using means and standard deviations, medians and interquartile ranges, or frequencies and proportions, as appropriate. All analyses were conducted using Python 3.10. Statistical and machine learning computations utilized: scikit-learn for preprocessing and evaluation metrics; XGBoost and LightGBM for binary classification; scikit-survival for survival analysis including Cox Proportional Hazards, Random Survival Forest, and Gradient Boosting Survival Analysis; SHAP for global explainability; LIME for local interpretability; and standard libraries including NumPy, pandas, matplotlib, and seaborn for data manipulation and visualization. Statistical significance was set at α = 0.05 for all hypothesis tests. All random seeds were fixed to ensure reproducibility.

### Data Preparation and Splitting

Data were randomly split into training (80%) and testing (20%) sets using stratified sampling based on the event indicator to maintain consistent event-to-censored ratios across both sets (12). This stratification was important for a balanced representation of the binary outcome (1-year mortality) in both datasets, which in turn is critical for reliable model evaluation in imbalanced clinical datasets (13). Random seed was set to 42 for reproducibility across all analyses.

### Missing Data Imputation

Missing data were handled using Multiple Imputation by Chained Equations (MICE), which is a robust iterative imputation approach that has demonstrated superior performance compared to simple imputation methods (14). Features were categorized as numerical (continuous or discrete with >10 unique values) or categorical (≤10 unique values). Numerical features were imputed using MICE with Random Forest regressors as the estimator, performing 10 iterations to achieve convergence (15). Random Forests were chosen as the imputation estimator due to their ability to capture non-linear relationships and interactions without requiring distributional assumptions (16). Categorical features were imputed using mode imputation. Critically, all imputation models were fit exclusively on training data and subsequently applied to the test set to prevent data leakage, which is important to avoid overly optimistic performance estimates (17).

### Machine Learning Model Development

Two gradient boosting algorithms were developed to predict binary 1-year mortality events. Extreme Gradient Boosting (XGBoost) was implemented using the XGBoost library, employing a regularized gradient boosting framework (18). Light Gradient Boosting Machine (LightGBM) was implemented as an advanced alternative, utilizing leaf-wise tree growth for enhanced efficiency (19). Both models underwent hyperparameter optimization using RandomizedSearchCV with 10-fold stratified cross-validation on the training set. The search space included: number of estimators (100, 200, 300), maximum depth (3–6), learning rate (0.01-0.1), subsample and feature sampling ratios (0.8-1.0), regularization parameters, and scale_pos_weight to address class imbalance. RandomizedSearchCV sampled 100 random combinations to balance computational efficiency with thorough exploration (20). Model performance was optimized using the area under the receiver operating characteristic curve (AUROC).

Time-to-event outcomes were analyzed using three complementary survival analysis methods appropriate for right-censored data (21). Time-to-event was measured in months from the index hospitalization. Random Survival Forest (RSF) extends Random Forests to survival outcomes by adapting tree-splitting criteria to maximize survival differences between nodes (22). Gradient Boosting Survival Analysis (GBSA) combines gradient boosting with Cox’s partial likelihood loss function (23). Cox Proportional Hazards regression was implemented as the benchmark clinical standard for time-to-event analysis (24). For the Cox model, features were standardized using StandardScaler, and L2 regularization (alpha=0.1) was applied to reduce overfitting (25,26). RSF and GBSA models underwent hyperparameter optimization using RandomizedSearchCV with 5-fold stratified cross-validation and concordance index (C-index) as the optimization metric (27). The search space for these models included: number of estimators (100–400), maximum depth, minimum samples per split and leaf, feature sampling strategies, and regularization parameters. All models were implemented using the scikit-survival package (28).

For feature selection, our primary approach used all candidate predictors with model-specific regularization (e.g., shrinkage/penalties; early stopping where applicable). As prespecified sensitivity analyses, we evaluated univariate pre-screening (for classifiers: F-test or mutual information; for survival: univariate Cox) to select top-k features before model fitting. All screening was performed within the CV loop (on training folds only) to avoid optimistic bias (11).

### Model Evaluation

Binary classification models were evaluated using AUROC, average precision, accuracy, balanced accuracy, F1-score, and confusion matrices on the held-out test set (29,30). Survival models were evaluated using the C-index, which measures the probability that the model correctly ranks predicted survival times for randomly selected patient pairs (27). C-index ranges from 0.5 (random) to 1.0 (perfect), with values >0.7 considered clinically useful (31). Time-dependent AUC was computed at multiple time points to assess discrimination as a function of follow-up time (32). Model calibration was assessed by comparing predicted survival probabilities to observed Kaplan-Meier estimates at 12 months using calibration plots (33). Inter-model agreement was assessed using Pearson correlation coefficients, and pairwise independent t-tests compared model performance (34). Bootstrap resampling with 1,000 iterations computed 95% confidence intervals for all performance metrics using the percentile method (35).

To compare AUROCs between classifiers on the same test set, we used the paired DeLong test (36). For differences in C-index between survival models, we used a paired, stratified bootstrap (1,000 replicates) to obtain ΔC with percentile CIs (35). We did not use independent t-tests for correlated performance measures.

### Model Explainability and Interpretability

SHapley Additive exPlanations (SHAP) provided global model interpretability by computing Shapley values from cooperative game theory (37). TreeExplainer was used for tree-based models, offering exact Shapley value computation (38). SHAP analyses included summary plots displaying feature importance and directional effects, bar plots showing mean absolute SHAP values, dependence plots illustrating feature relationships and interactions, and waterfall plots for individual predictions. Local Interpretable Model-agnostic Explanations (LIME) provided complementary instance-level explanations by fitting local linear approximations around individual predictions (39). LIME explanations were generated for representative patients across the risk spectrum (high-risk, median-risk, low-risk) using 15 features and 5,000 perturbations per instance. For survival models lacking native feature importance metrics, permutation importance was computed by measuring the decrease in C-index after randomly shuffling each feature (40,41).

## RESULTS

### Cohort and Outcome

From 2010–2020, we identified 7,840 unique first admissions for HF exacerbation among patients with HFpEF on admission echocardiography. Mean age was 78 years (IQR 68–86); 4,362 (55.6%) were women, and 7,408 (94.5%) self-identified as White. Hypertension was common (86.1%), followed by hyperlipidemia (71.8%) and diabetes (53.9%). Among those with available data, 76.3% had RVSP ≥35 mmHg. Median admission laboratory values were: albumin 3.6 g/dL (IQR 3.2–3.9), NT-proBNP 2,391 pg/mL (IQR 964–5,169), and eGFR 55 mL/min/1.73 m² (IQR 37–74.5). Baseline characteristics, including demographics, comorbidities, and medications, are provided in Table 1, while echocardiographic and lab findings are shown in Supplementary Tables S1 and S2, respectively. During a 1-year follow-up, 2,466 deaths (31.5%) occurred. Data were randomly split into training (n=6,272; 80%) and test (n=1,568; 20%) sets with similar mortality (31.4% vs 31.6%).

**Table 1.** Baseline Demographics, Comorbidities, and Medications at Discharge.

|  |  |
| --- | --- |
| Demographics | N (%) |
| Sex |  |
| Female | 4362 (55.6%) |
| Male | 3478 (44.4%) |
| Race |  |
| Asian | 60 (0.8%) |
| Black | 169 (2.2%) |
| Native American | 24 (0.3%) |
| White | 7408 (94.5%) |
| Other | 85 (1.1%) |
| Unknown | 94 (1.2%) |
| Age (years) |  |
| < 50 | 312 (4.0%) |
| 50-64 | 1142 (14.6%) |
| 65-79 | 2834 (36.1%) |
| 80-94 | 3273 (41.7%) |
| > 94 | 279 (3.6%) |
| Body Mass Index (Kg/m<sup>2</sup>) |  |
| < 25 | 1811 (23.1%) |
| 25.0-29.9 | 2023 (25.9%) |
| 30.0-34.9 | 1587 (20.3%) |
| 35.0-39.9 | 1041 (13.3%) |
| ≥ 40 | 1362 (17.4%) |
| Missing | 16 |
| Comorbidities |  |
| CAD | 1635 (20.9%) |
| COPD | 3058 (39.0%) |
| DM | 4223 (53.9%) |
| HTN | 6754 (86.1%) |
| Hyperlipidemia | 5632 (71.8%) |
| TIA | 2112 (26.9%) |
| Medications at Discharge |  |
| Aldosterone Antagonists | 363 (4.9%) |
| Diuretics | 4008 (51.1%) |
| SGLT2 Inhibitors | 7 (0.1%) |
| CAD: Coronary Artery Disease, COPD: Chronic Obstructive Pulmonary Disease, DM: Diabetes Mellitus, HTN: Hypertension, PF: Pulmonary Fibrosis, TIA: Transient Ischemic Attack |  |

### Predictor Screening

Of 51 candidate predictors, 39 (76.5%) were associated with 1-year mortality by F-test. The strongest signals included albumin, age, NT-proBNP, renal failure, BUN, body mass index (BMI), hemoglobin/hematocrit, and left ventricular end-diastolic dimension (LVEDD). Supplemental Table S3 shows the univariate Cox model output, each variable’s coefficients, and effect estimates, while Supplemental Table S4 shows the multivariate Cox model output.

### Binary Event Prediction (XGBoost, LightGBM)

Both boosted classifiers showed moderate discrimination on the held-out test set with overlapping CIs: XGBoost AUROC 0.751 (95% CI 0.727–0.775); AP 0.575 (95% CI 0.529–0.624) and LightGBM AUROC 0.749 (95% CI 0.721–0.776); AP 0.577 (95% CI 0.532–0.625). The AP values were well above the event-rate baseline (0.315). Discrimination and calibration are shown in Figure 1 (ROC and PR curves) and Supplemental Figure S1 (calibration); calibration was close to the 45° line with mild under-prediction at very low risk. Predicted probabilities from the two models were highly concordant (r = 0.962; Supplemental Figure S1).

**Figure 1:**
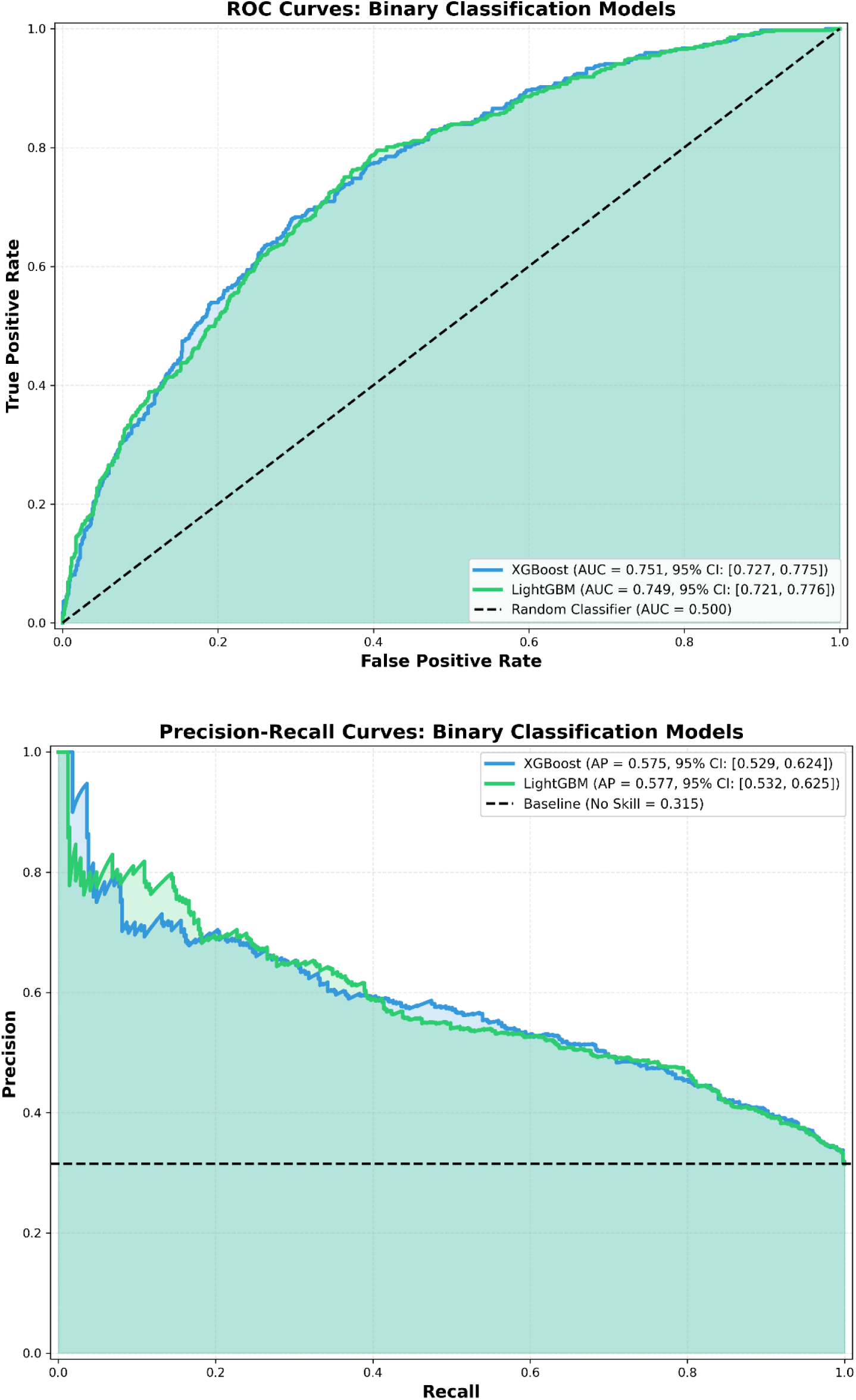
ROC (upper panel) and PR (lower panel) curves for the two binary classification models (XGBoost and LightGBM). Receiver operating characteristic (ROC) curves (Panel A) and precision-recall (PR) curves (Panel B) comparing XGBoost and LightGBM classifiers on the held-out test set (n=1,568). Both models demonstrated moderate discrimination with overlapping performance: XGBoost achieved an area under the ROC curve (AUROC) of 0.751 (95% CI: 0.727–0.775) and LightGBM 0.749 (95% CI: 0.721–0.776). Average precision (AP) exceeded the no-skill baseline (0.315), confirming utility beyond class prevalence. The dashed diagonal line (Panel A) represents random classification (AUROC=0.50); the horizontal dashed line (Panel B) represents the event-rate baseline. Shaded regions indicate 95% confidence intervals. CI, confidence interval.

Regularized all-feature models outperformed univariate pre-screened variants (e.g., XGBoost-MI-25; LightGBM-F-test-25), so the regularized models are primary. A paired bootstrap comparison of AUROCs yielded a very small mean difference (∼0.002) with a 95% CI spanning zero; we therefore interpret no clear performance difference (Supplement Table S5).

### Time-to-Event Prediction (Cox, RSF, GBS)

Among survival models, gradient boosting survival (GBS) had the highest discrimination, followed by random survival forest (RSF) and Cox Elastic Net (Figure 2): GBS C-index 0.718; RSF 0.711; Cox 0.704. Pairwise bootstrap comparisons favored GBS > RSF (ΔC 0.0062, 95% CI 0.0052–0.0071) and RSF > Cox (ΔC 0.0076, 95% CI 0.0065–0.0085); GBS > Cox (ΔC 0.0137, 95% CI 0.0127–0.0147) (Supplement Table S6). Calibration-at-12-months plot showed good agreement for all three models (Figure S2). Pre-selection to a reduced feature set lowered performance for RSF/GBS, so all-feature fits are primary.

**Figure 2:**
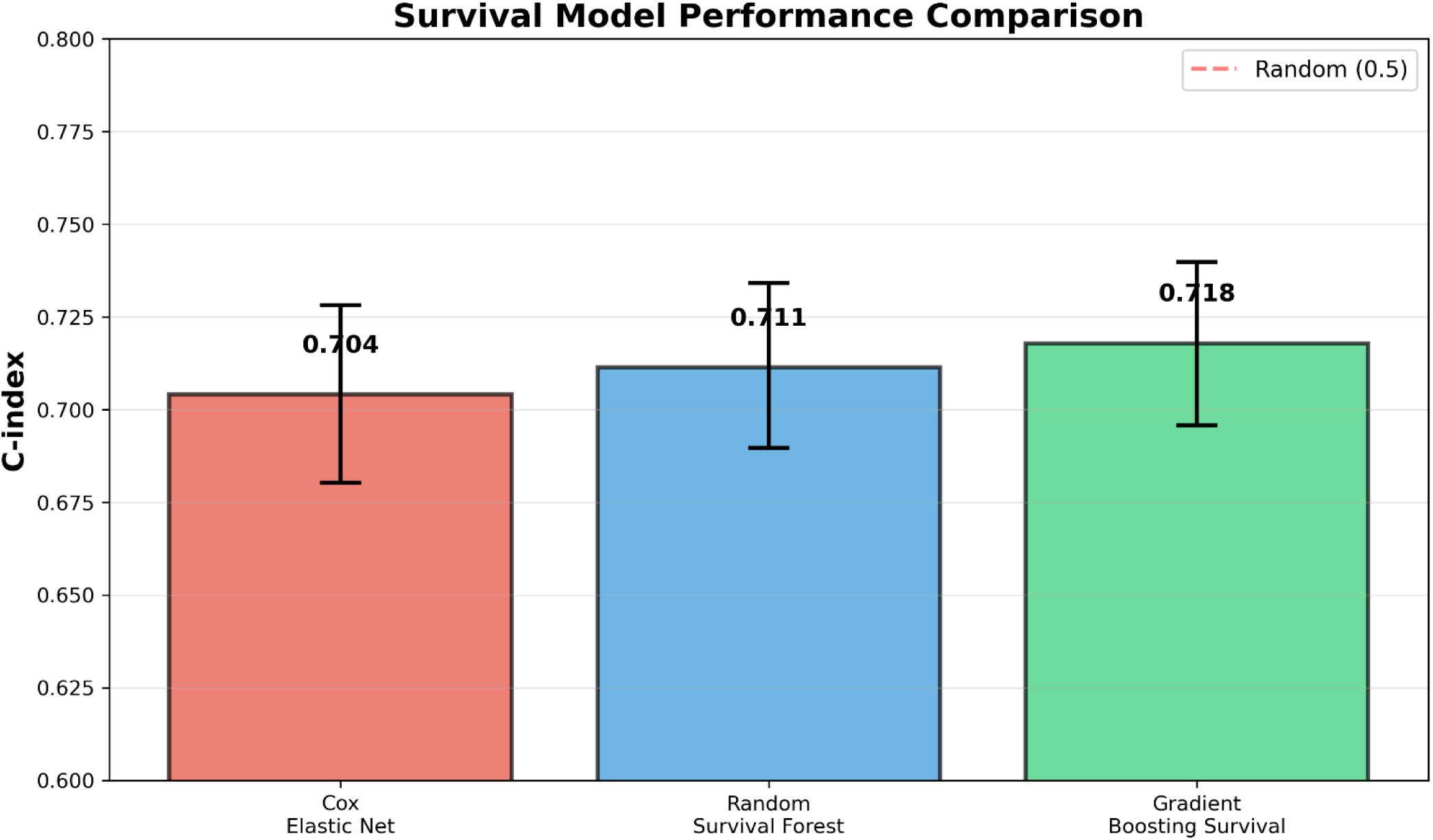
C-index comparison (time-to-event prediction) for the Cox Elastic Net, Random Survival Forest (RSF), and Gradient Boosting Survival (GBS) models. Bar plot comparing concordance index (C-index) across three time-to-event models evaluated on the test set. Gradient Boosting Survival (GBS) achieved the highest discrimination (C-index=0.718), followed by Random Survival Forest (RSF; 0.711) and Cox Elastic Net (0.704). Error bars represent bootstrap 95% confidence intervals (1,000 iterations). Pairwise comparisons confirmed statistically significant differences (GBS vs. Cox: ΔC=0.014). All models exceeded random prediction (C-index=0.50, indicated by red dashed line), demonstrating clinically useful prognostic accuracy for one-year mortality prediction in heart failure with preserved ejection fraction.

### Independent Risk and Protective Factors (Multivariable Cox)

In the fully adjusted model, several variables remained independently associated with higher 1-year mortality (Figure 4 and Supplementary Table S3). The strongest associations were observed for hematocrit (HR 1.41, 95% CI 1.16–1.77), age (HR 1.39, 95% CI 1.30–1.49), renal failure (HR 1.19, 95% CI 1.13–1.26), BUN (HR 1.19, 95% CI 1.12–1.27), and NT-proBNP (HR 1.11, 95% CI 1.05–1.17).

Conversely, several variables were independently protective after adjustment (Figure 4 and Supplementary Table S3). Higher hemoglobin (HR 0.63, 95% CI 0.51–0.76) and higher albumin (HR 0.70, 95% CI 0.66–0.74) were associated with lower risk, consistent with the explainability analyses. SGLT2-inhibitor use was also associated with lower 1-year mortality (HR 0.73, 95% CI 0.72–0.78). Additional independent protective associations included LVEDD (HR 0.80, 95% CI 0.67–0.91), higher BMI (HR 0.84, 95% CI 0.78–0.90), higher systolic blood pressures at time of echo (SBP_echo; HR 0.86, 95% CI 0.82–0.91), and platelet count (HR 0.87, 95% CI 0.82–0.92).

### Signal Coherence Across Approaches

Across classification and survival frameworks, the leading predictors were consistent: lower albumin, older age, higher NT-proBNP, renal dysfunction markers (renal failure/BUN), lower hemoglobin/hematocrit, and larger LVEDD were associated with higher risk; SGLT2-inhibitor use was associated with lower risk in univariate survival analysis. (Figure 3, Supplemental Figures 3-6) These patterns align with clinical expectations for HF/HFpEF and informed interpretation.

**Figure 3:**
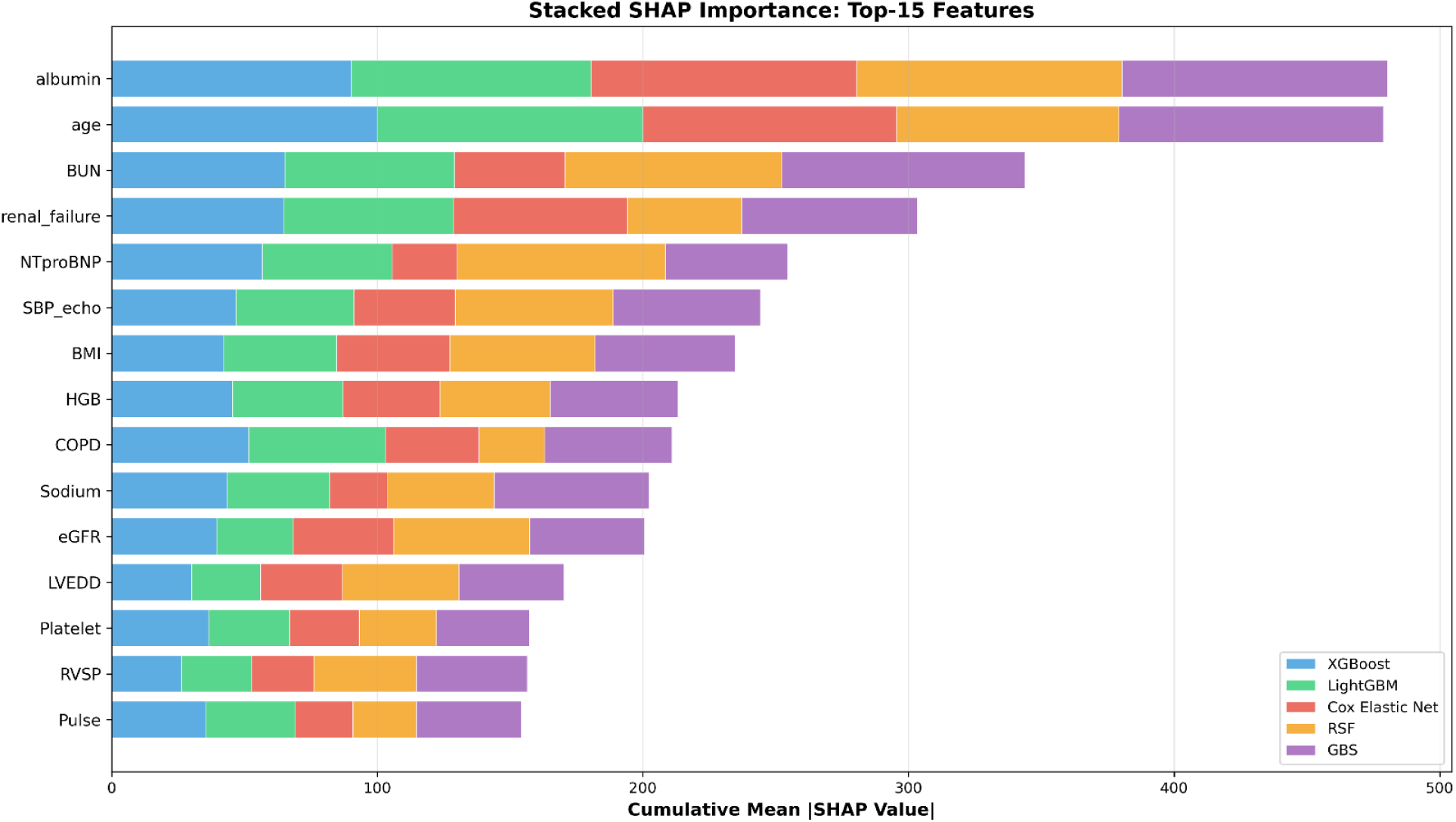
Stacked SHAP importance (top-15). Stacked horizontal bar plot displaying cumulative mean absolute SHapley Additive exPlanations (SHAP) values for the top-15 predictors across all five models. Albumin and age contributed the largest share of predictive importance, followed by blood urea nitrogen (BUN), renal failure, and N-terminal pro-B-type natriuretic peptide (NTproBNP). Color segments represent individual model contributions: XGBoost (teal), LightGBM (green), Cox Elastic Net (coral), Random Survival Forest (RSF; purple), and Gradient Boosting Survival (GBS; gold). The consistency of rankings across algorithm families demonstrates signal coherence and supports clinical interpretability. SBP_echo, systolic blood pressure at echocardiography; HGB, hemoglobin; eGFR, estimated glomerular filtration rate; LVEDD, left ventricular end-diastolic dimension; RVSP, right ventricular systolic pressure.

**Figure 4:**
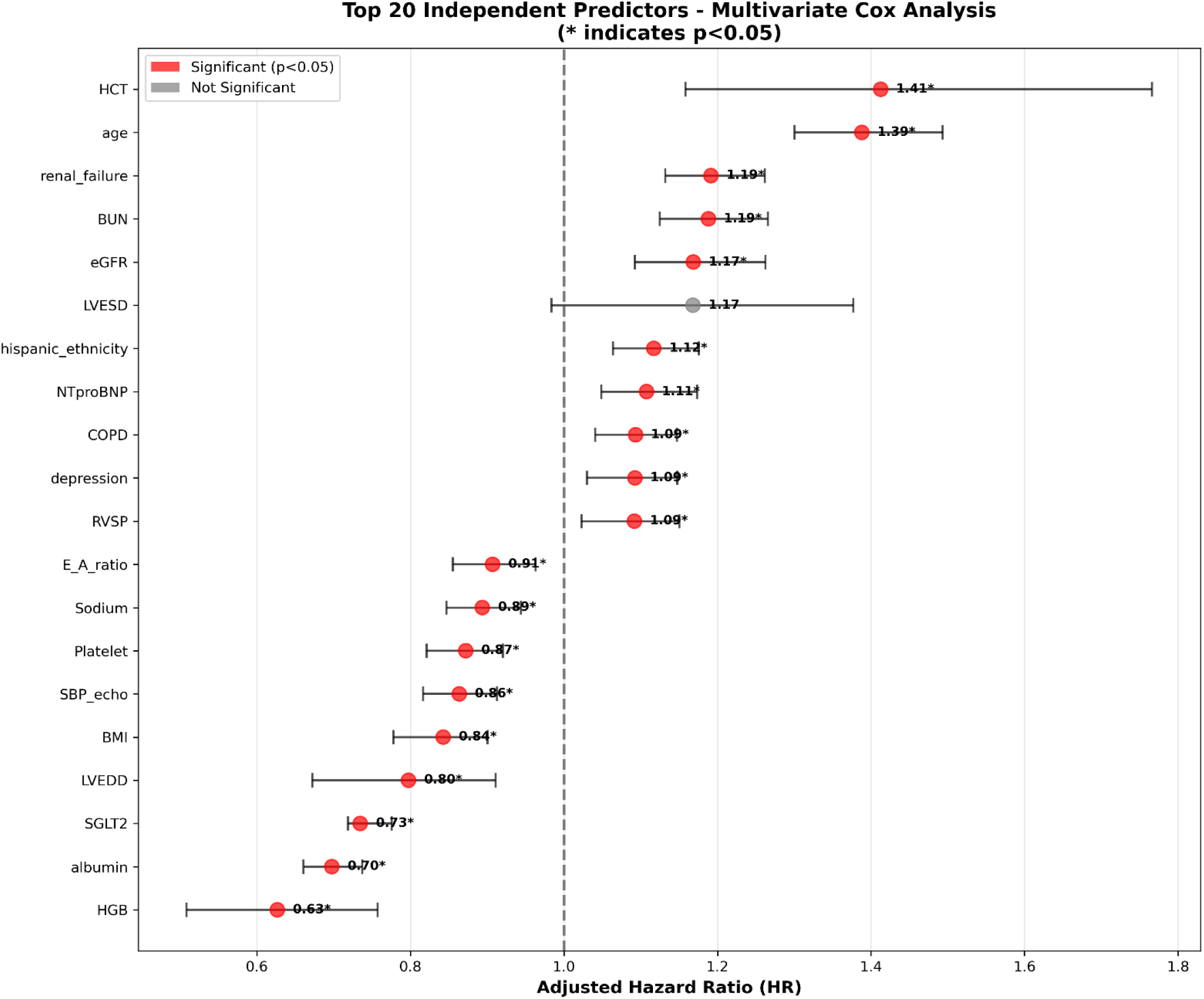
Top 20 multivariate predictors and the HRs (95% CIs) for each predictor. Forest plot displaying adjusted hazard ratios (HRs) with 95% confidence intervals for the top 20 independent predictors of one-year mortality. Red circles indicate statistically significant associations (p<0.05); asterisks denote significance. The vertical dashed line represents HR=1.0 (no effect). Higher hematocrit (HCT; HR=1.41), age (HR=1.39), and renal failure (HR=1.19) were associated with increased mortality, while higher hemoglobin (HGB; HR=0.63), albumin (HR=0.70), and SGLT2 inhibitor use (HR=0.73) were protective. Horizontal lines represent 95% CIs. BUN, blood urea nitrogen; eGFR, estimated glomerular filtration rate; LVESD, left ventricular end-systolic dimension; NTproBNP, N-terminal pro-B-type natriuretic peptide; RVSP, right ventricular systolic pressure; E_A_ratio, mitral E/A ratio; SBP_echo, systolic blood pressure at echocardiography; BMI, body mass index; LVEDD, left ventricular end-diastolic dimension.

### Model Explainability

#### Global patterns and cross-model stability

Per-model SHAP summary plots identified a stable set of high-impact predictors with consistent directionality (Figure 3, Supplemental Figures S4-12): albumin and age were top-ranked across all models, followed by BUN, renal failure, NT-proBNP, SBP_echo, BMI, hemoglobin (HGB), eGFR, sodium levels, and LVEDD. Color gradients showed expected directions: older age and higher NT-proBNP/BUN pushed predictions upward; lower albumin and lower eGFR also pushed upward; higher SBP_echo and higher hemoglobin tended to be protective.

Two cross-model visual summaries emphasize stability. The radar plot of the top-8 features (normalized 0–100) shows albumin levels and age near 100 for every model; survival tree/boosting models place relatively higher weight on BUN (highest in GBS) and NT-proBNP (highest in RSF), while penalized Cox down-weights these relative to trees (Supplemental Figure S5). The stacked top-15 plot shows the cumulative contribution across models; albumin + age account for the largest share, followed by BUN, renal failure, NT-proBNP; the remainder form a longer tail (Figure 3).

The SHAP comprehensive statistics (Supplemental Table S7) confirm the average ranking for each variable: albumin, age, BUN, NT-proBNP, SBP_echo, BMI, and HGB. This feature consistency shows that each of these appeared in the top-20 of all five models (Supplemental Figure S7-12) with favorable average ranks. Inter-model agreement was high in the model-agreement heatmap (Supplementary Figure S6): SHAP-importance correlations were 0.994 (XGBoost–LightGBM) and 0.913 (RSF–GBS); the lowest pair, Cox–RSF, remained substantial (0.775).

#### Local, patient-level explanations

LIME panels illustrate how predictors combine within individuals and mirror the global patterns (Supplementary Figures S13-17). In high-risk profiles, the largest positive weights concentrated on very old age (>86 yrs), low albumin, high BUN, renal failure, and very high NT-proBNP; median-risk cases showed mixed contributions (e.g., low-normal albumin and renal impairment partly offset by other features); low-risk cases were characterized by preserved renal function (eGFR ≳ 74ml/min/1.73m^2^), higher albumin (>3.2gm/dl), and SGLT2 therapy with low BUN and absence of right ventricular (RV) abnormalities. The LIME summary table (Supplemental Table S8) aggregates these patterns across the binary models, as an example, and patient risk strata.

## DISCUSSION

In a contemporary HFpEF cohort, gradient-boosted classifiers achieved moderate discrimination for 1-year mortality (AUROC ≈0.75) with acceptable calibration, while among survival models, gradient boosting survival performed best (C-index 0.718), followed by random survival forest and penalized Cox. Across algorithm families, the same clinical signals dominated risk: lower albumin (<3.2gm/dl), older age, higher NT-proBNP, and renal dysfunction (BUN/renal failure) consistently pointed to higher risk; SGLT2 inhibitor use was associated with lower risk in univariate survival analysis. These findings align with established epidemiology showing HFpEF in older, frequently female patients, with rising prevalence tied to population aging and cardiometabolic multimorbidity (42–45).

Given the public health significance of HFpEF and its steadily increasing prevalence due to the aging of the population and accumulation of risk factors, establishing prognostic models in a real-life patient population has direct implications for clinical care. These current models validate our previous work (46) in a significantly larger, somewhat more diverse patient population. Our results extend prior HFpEF risk work by showing that modern tree/boosting methods and Cox largely agree on what matters, rather than uncovering new, model-specific signals. The repeated prominence of albumin and age across all models reinforces the prognostic value of nutritional/inflammatory status and biologic aging/frailty in HFpEF outcomes (42,43,47,48). In this study, an albumin level of <3.2 g/dL remained the single highest risk factor as both an independent predictor of prognosis with a hazard ratio (HR) of 2.006 (95% CI: 1.818-2.200) and as a multivariate predictor in both models including and excluding NT-proBNP (HRs of 1.713 [95% CI 1.535-1.920]and 1.733 [95% CI 1.551-1.915], respectively). Low albumin levels are being increasingly recognized as an adverse prognosticator in the setting of HFpEF by multiple investigators (49,50), correlating with longer hospital stays in males hospitalized for HF exacerbation (51), and also has been associated with the development of HF in adults with baseline low levels of albumin 10 years post follow-up (52). Furthermore, low albumin levels remained a strong predictor of the primary endpoint in the TOPCAT trial even after adjustment for the MAGGIC risk score (HR 0.72, 95% CI 0.67-0.78; p <0.0001)(48). Similarly, in another cohort, albumin enhanced risk stratification beyond NT-proBNP and the MAGGIC score (adjusted standardized HR 0.56, 95% CI 0.37-0.83)(50). The role of chronic systemic inflammation in the pathophysiology of HFpEF is being realized increasingly(53). Sustained elevation in TNFα and IL6 levels suppress hepatic albumin production and increase capillary permeability, promoting albumin leakage into the interstitial space and urine(54,55), hence linking inflammation directly with low albumin in HFpEF and consequently adverse prognosis.

NT-proBNP has prognostic value despite known variability and multi-system influences, (5,56,57) supporting its use as part of a cluster of markers rather than a stand-alone gatekeeper. Increased NT-proBNP levels are often associated with both the incidence and poor prognosis of HFpEF (58,59), a finding also supported by our data. After multivariable adjustment, NT-proBNP remained independently associated with higher 1-year mortality (adjusted HR 1.11, 95% CI 1.05–1.17). NT-proBNP’s value as a prognostic marker, however, is tempered by its susceptibility to elevation in multiple conditions, including peripheral artery disease (60), acute ischemic stroke (61), hypertension (62), COPD (63), atrial fibrillation (64), and more (65). NT-proBNP has been criticized as a prognosticator for HF in the past due to the obvious abundance of conditions that can cause an elevation in these levels, as well as extremely large variations between patients (5). Indeed, in our study, NT-proBNP ranged from 5 to 65,900 pg/mL (mean 4,476.8 pg/mL, SD 6,240.2). An NT-proBNP ≥5,000 pg/mL corresponded to a materially higher unadjusted risk (≈22% relative increase). The direction of association for SGLT2 inhibitors is consistent with randomized evidence in HFpEF/mid-range EF populations (66,67); we do not infer causality from observational signal but note concordance with well-established trial data.

Explainability was prespecified. Global SHAP analyses showed high cross-model agreement (e.g., XGBoost-LightGBM r ≈ 0.99; RSF-GBS r ≈ 0.91), with albumin and age repeatedly top-ranked. Local (LIME) panels produced patient-level narratives that mirrored the global signals: very old age, low albumin, renal dysfunction, and markedly elevated NT-proBNP in high-risk profiles; preserved renal function, higher albumin, and SGLT2 therapy in low-risk profiles (37,39). These tools make the models auditable and clinically interpretable, while we acknowledge that post-hoc explanations quantify associations within a fitted model and are not causal (68,69).

Two practical messages emerge. First, risk is driven by a recurrent cluster (albumin, age, renal indices, NT-proBNP) of markers easily available on admission laboratory values. This supports routine risk stratification and targeted actions (nutrition support, renal-protective strategies, guideline-directed therapy, and SGLT2 eligibility checks) aligned with individual risk profiles (42,66,67). Second, because discrimination and calibration are somewhat similar across methods, health systems can adopt either a boosted classifier (for 1-year probability) or GBS/RSF (for time-to-event estimates) and focus operational effort on decision thresholds and net benefit (70,71). Consistent with reporting guidance, any deployment should include calibration monitoring and updating (11).

### Limitations

Our study has several limitations worth noting. This is an internally validated study from a predominantly Caucasian cohort; external validation across health systems and demographics is needed (11). We relied on routinely collected data subject to coding and measurement error. We did not model cause-specific mortality or competing risks. Although explainability improved transparency, SHAP/LIME do not imply causation, and treatment decisions should continue to rely on trial-proven therapies (66,67,69). Future prospective evaluation with decision-curve analysis, and pre-specified decision-thresholds is needed to define clinical utility and implementation logistics (70,71). Adding calibration intercept/slope and Brier-based metrics to routine monitoring, exploring time-updated features (labs/vitals trajectories), and testing workflow integrations (EHR alerts, risk-stratified follow-up) are practical next steps. Subgroup performance and equity assessments should be reported alongside deployment. Finally, explainability can be extended to care-pathway audits (e.g., mapping low albumin and renal dysfunction to nutrition consults and renal-protective strategies) to close the loop from prediction to action.

### Conclusion

Across five modeling families, we observed convergent, clinically coherent predictors of 1-year mortality in HFpEF and consistent performance with acceptable calibration. The dominance of albumin, age, renal indices, and NT-proBNP, and the agreement of model-agnostic explanations, suggest that a transparent, implementable risk tool is feasible using data available at admission. External validation and prospective testing should precede routine use, but these results outline a clear, interpretable foundation for risk-guided HFpEF care.

## Data Availability

All data is presented in the manuscript and supplemental material. Original, deidentified, raw data is available from the authors upon request.

## Notes

**Funding:** JL and MZ are supported by NIH grant R01HL153407 awarded to MZ.

### Competing Interest Statement

The authors have declared no competing interest.

### Clinical Trial

Not applicable

### Funding Statement

JL and MZ are supported by NIH grant R01HL153407 awarded to MZ.

### Author Declarations

Mayo Clinic, Rochester, IRB

